# SARS-CoV-2 On-the-Spot Virus Detection Directly from Patients

**DOI:** 10.1101/2020.04.22.20072389

**Authors:** Nadav Ben-Assa, Rawi Naddaf, Tal Gefen, Tal Capucha, Haitham Hajjo, Noa Mandelbaum, Lilach Elbaum, Peter Rogov, King A. Daniel, Shai Kaplan, Assaf Rotem, Michal Chowers, Moran Szwarcwort-Cohen, Mical Paul, Naama Geva-Zatorsky

## Abstract

Many countries are currently in a lockdown state due to the SARS-CoV-2 pandemic. One key aspect to transition safely out of lockdown is to continuously test the population for infected subjects. Currently, detection is performed at points of care using quantitative reverse-transcription PCR (RT-qPCR), and requires dedicated professionals and equipment. Here, we developed a protocol based on Reverse Transcribed Loop-Mediated Isothermal Amplification (RT-LAMP) for detection of SARS-CoV-2. This protocol is applied directly on SARS-CoV-2 nose and throat swabs, with no RNA purification step required. We tested this protocol on over 180 suspected patients, and compared its results to the standard method. We further succeeded to apply the protocol on self-sampled saliva from confirmed cases. Since the proposed protocol provides results on-the-spot, and can detect SARS-CoV-2 from saliva, it can allow simple and continuous surveillance of the community.

## Introduction

During a pandemic, surveillance is crucial for minimizing viral spread. The common and approved detection method worldwide requires professional experience in sampling, performing the reaction and analyzing the results. Moreover, it requires dedicated machines, and chemical reagents as well as sophisticated sample collection and transport logistics. Due to these laborious and cumbersome requirements, the number of detection tests per day is limited, and many patients in the community are not sampled, let alone sampled frequently. Such limited surveillance necessitates global and strict quarantine requirements, which threaten the global economy.

Detection is key. We believe that a simple and easy detection method, preferably one that can be performed and interpreted on-the-spot could relieve some of the current limitations, and help execute an efficient and safe exit strategy from lockdowns. Fast and simple serological tests that can, in principle, be applied in households, are being developed ^[1]^. However, anti-viral antibodies can be detected only several days after the infection onset, and can persist even after clearance of the virus. A stage at which the patient is not contagious anymore ^[2]^. Hence, the presence of antibodies detected in such home-kits are an indirect indication on previous viral exposure. These tests do not account to the actual viral load—a critical parameter for minimizing the spread.

Detection of viral nucleic acids in patients is the gold standard detection method to date. It is currently performed at hospitals by professionals. As opposed to antibodies, detection of the viral RNA is a direct measure for the contagiousness of the patient. At this stage of the current COVID-19 pandemic, it is clear that the availability and throughput of standard methods for viral nucleic acid detection is limited both by resources and accessibility to the community.

Standard detection methods for viral RNA in patients include RNA purification, reverse transcription and quantitative PCR (RT-qPCR). These processes are time consuming, require multiple biochemical reagents, lab-grade instruments and trained professionals ^[3]^. Fortunately, to date, alternative molecular biology methods can overcome these limitations. One of these methods is colorimetric Loop-Mediated Isothermal Amplification (LAMP) ^[4]^. LAMP is performed at a single and constant temperature allows a one-step reverse transcription and its results can be visualized by color change (for graphical presentation see Fig. 1a). Due to the need for reverse transcription, this method is called reverse-transcribed (RT)-LAMP. Altogether, these advantages eliminate the need for sophisticated lab equipment ^[5-7]^. Here, we adjusted RT-LAMP for the detection of SARS-CoV-2 RNA directly from clinical diagnostic swabs of human patients, without RNA purification steps. The primers we used were previously designed and validated by Zhang *et al*. ^[4]^ (see primers table). We studied samples from positive and negative patients for the SARS-CoV-2. The samples were confirmed by approved RNA purification and quantification at the Rambam Health Care Campus (RHCC) hospital.

**Fig. 1:**
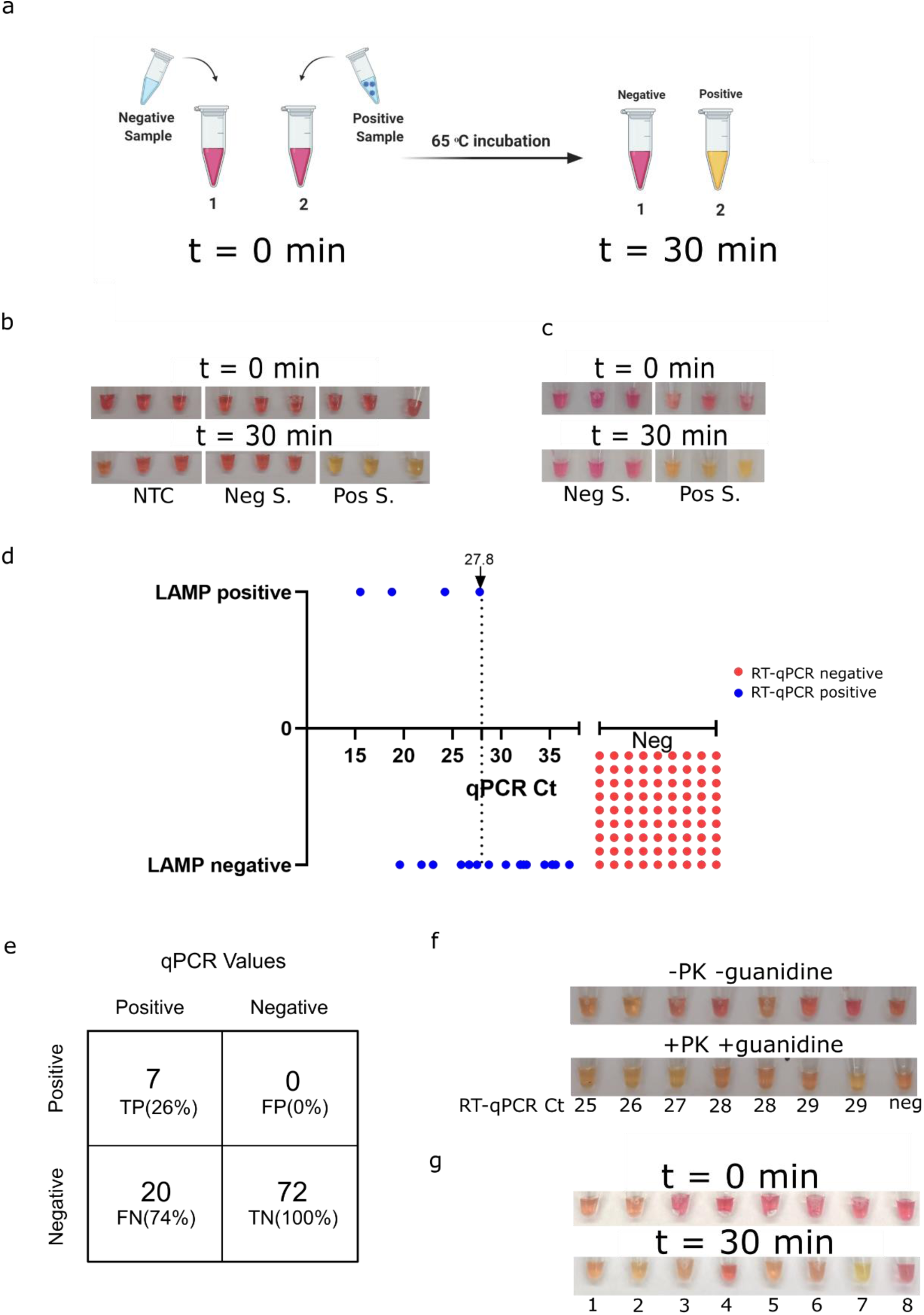
Protocol adjustment and optimal conditions. **a**, Schematic representation of the isothermal, colorimetric RT-LAMP reaction. **b**, RT-LAMP reaction on purified RNA from nasal and throat swabs submerged in UTM buffer. Results shown at t=0 and after 30 minutes incubation at 65° C. (left) No template control (NTC), (middle) negative subject (Neg S.) and (right) positive subject (Pos S.). Three technical replicates of each sample are shown. **c**, Representative pictures of RT-LAMP test results of clinical diagnostic nasal and throat swabs. Samples were directly tested with no RNA purification step. (left) three different negative samples (Neg S.), and (right) three different positive samples (Pos S.) at t = 0 and t = 30 minutes. **d**, Comparison of the RT-LAMP method to the Ct values of the standard RT-qPCR results (3 true positive and 2 false negative samples out of the 99 that were analyzed, are not shown due to inaccessibility to their RT-qPCR Ct values). RT-qPCR negative samples were assigned arbitrary Ct values, for visualization. **e**, Classification of true positive (TP), true negative (TN), false positive (FP) and false negative (FN) numbers and rate of RT-LAMP test results in comparison to the standard RT-qPCR test results. **f**, Clinical diagnostic nasal and throat swabs tested by two different RT-LAMP protocols. Upper panel, without proteinase K and guanidine hydrochloride. Lower panel, with proteinase K treatment and guanidine hydrochloride. For RT-qPCR positive samples, Ct value is presented under each sample, the sample to the right is negative. **g**, RT-LAMP results on samples from patients confirmed to be positive for the following virus: 1-2, HSV swabs (lysed and inactivated as described in the currently developed protocol). 3, HSV purified DNA. 4, RSV purified RNA. 5, Influenza B RNA. 6, Enterovirus RNA. 7, RNA extraction from SARS-CoV-2 positive patient. 8, no template control. Results are shown at t = 0 and t = 30 minutes after incubation at 65º C.

## Results

### Optimal conditions of colorimetric RT-LAMP SARS-CoV-2 detection

We first applied RT-LAMP on purified RNA from COVID-19 positive and negative swabs. As in Zhang *et. al*., ^[4]^ the RT-LAMP results agreed with the standard RT-qPCR results (Fig. 1b). To simplify the detection method, we sought to test the RT-LAMP reaction on clinical diagnostic throat and nose swabs from patients. These swabs were kept in universal transfer media (UTM). To these samples, we added an inactivation step by heating the UTM to 95°C for 5 minutes. Inactivated samples from confirmed patients were found to be positive by the RT-LAMP reaction (Fig. 1c). We then evaluated this RT-LAMP protocol on a cohort of 99 patients that were tested at the hospital. This pool included 27 positive samples with a wide range of viral load and 72 negative samples. Samples were previously evaluated by the standard RT-qPCR test. The detection limit of this RT-LAMP protocol was at cycle threshold (Ct) of 27.8 (Fig. 1d, Fig. S1 and Supplementary Table 1), with 7 true positives (TP), 20 false negatives (FN), 72 true negatives (TN), and no false positives (FP) (Fig. 1e, Fig. S1 and Supplementary Table 1). Although there were no false positives, the rate of TP was very low. Hence, we were interested to improve the RT-LAMP protocol of detection from clinical diagnostic patient swabs. Since these swab samples may contain enzymatic inhibitors that might affect the efficiency of viral RNA detection, we have tested the addition of proteinase K and guanidine hydrochloride to the process. Proteinase K was added to the UTM sample taken from the original tube, and guanidine hydrochloride to the RT-LAMP reaction step. We first compared the two protocols on eight patients with low, medium and high Cts. Out of seven patients with positive results of RT-qPCR (Ct<37), two were clearly RT-LAMP positive in the former protocol and four in the new protocol. We concluded that these adjustments improved the RT-LAMP efficiency of viral RNA detection directly from clinical diagnostic swab samples (fig. 1f). Lastly, we tested samples from patients confirmed to be infected with viruses other than SARS-CoV-2. We tested a few patients that were previously confirmed to be infected by other viruses (HSV, RSV, Influenza and Enterovirus). As shown in figure 1g, this RT-LAMP reaction was negative for patients infected with these viruses, and positive for SARS-CoV-2.

### Adjusted protocol on cohort of 83 suspected patients

With this adjusted protocol, we set to validate it on an additional cohort of 83 patients suspected of SARS-CoV-2. These patients were tested at RHCC by the standard RT-qPCR, 31 were negative and 52 were positive with a wide range of Ct values (14-35). We were interested in finding the optimal incubation time to yield the best rate of true positives without increasing the rate of false negatives. We performed the RT-LAMP reaction for up to 40 minutes, and evaluated the colorimetric results at time-points 30, 35 and 40 minutes. With time, the number of TP increased, the number of FN decreased, with no change in the numbers of TN and with one FP throughout (Fig. 2a, b, Fig. S2 and Supplementary Table 2). Time-points 35 and 40 showed the highest TP rate in samples with low (Ct<26) and medium (26<Ct<29) Ct values (Fig. 2d, Fig. Supplementary Table 2). Hence, in these conditions, time-points 35 and 40 were better altogether. Figure 2c shows the results of the adjusted method at time point 40. These results were compared to the RT-qPCR Ct values of the same patients. RT-LAMP was most sensitive in detection of positive patients with viral load that corresponds to low and medium Ct values. Under Ct 28.8, the true positive rate of RT-LAMP was 93%. Test results were interpreted without knowledge of the results of the reference standard.

**Fig. 2:**
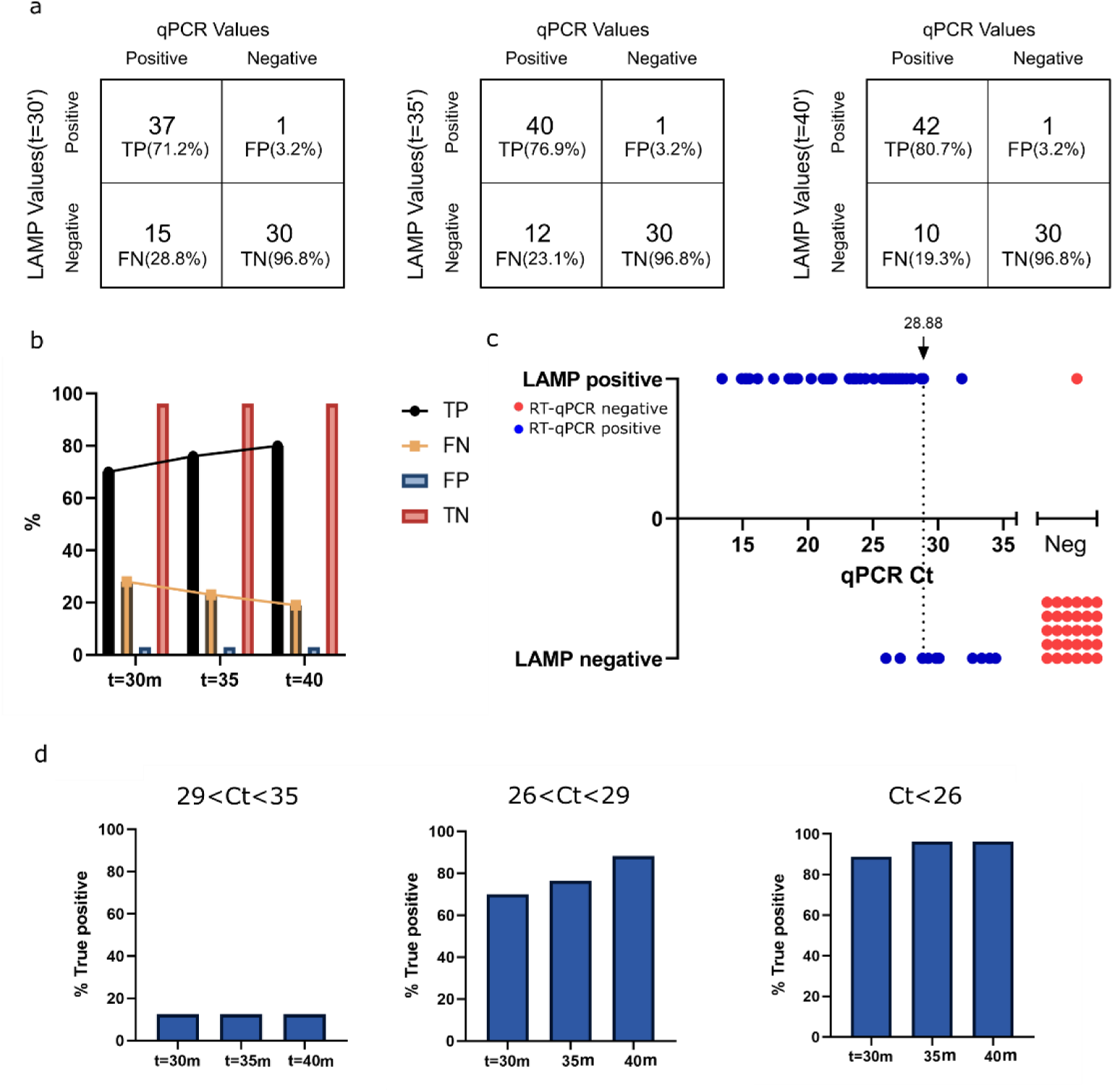
Adjusted RT-LAMP protocol tested on 83 clinical diagnostic nasal and throat swab samples. **a**, Classification of true positive (TP), true negative (TN), false positive (FP) and false negative (FN) numbers and rates of RT-LAMP test results in comparison to the standard RT-qPCR results. Boxes from left to right represent results at t = 30, 35 and 40 minutes, respectively. **b**, bar graph representation of the TP, TN, FP and FN rates shown in (a). **c**, Comparison of the RT-LAMP method to the Ct values of the standard RT-qPCR. **d**, Graphical representation of TP rates of RT-LAMP in different incubation periods, t = 30 min, t = 35 min, t = 40 min, compared to RT-qPCR test results separated by Ct value intervals. 29<Ct<35 (left), 26<Ct<29, (middle), Ct<26 (right).

### Applying RT-LAMP protocol on saliva samples from confirmed patients

Human to human transmission of SARS-CoV-2 is mainly through saliva droplets ^[8]^. A comparison between saliva samples to the standard swabs showed a higher viral load in the saliva ^[9]^. Moreover, the FDA has recently approved saliva as a possible way of sampling for COVID-19 (https://www.fda.gov/media/136875/download). Therefore, we next performed RT-LAMP on human saliva samples. Saliva samples were self-collected from three different confirmed patients, and one suspected negative subject. In parallel, we also performed the standard RNA purification and RT-qPCR reactions in the hospital setting. The confirmed patients were found positive in both RT-LAMP and RT-qPCR from saliva, and the suspected negative subject was confirmed negative (Fig. 3a). Here, as a positive control for the reaction and saliva sampling, we tested for the human gene POP7, in addition to gene N of SARS-CoV-2. POP7 was detected in all saliva samples (Fig. 3a).

**Fig. 3:**
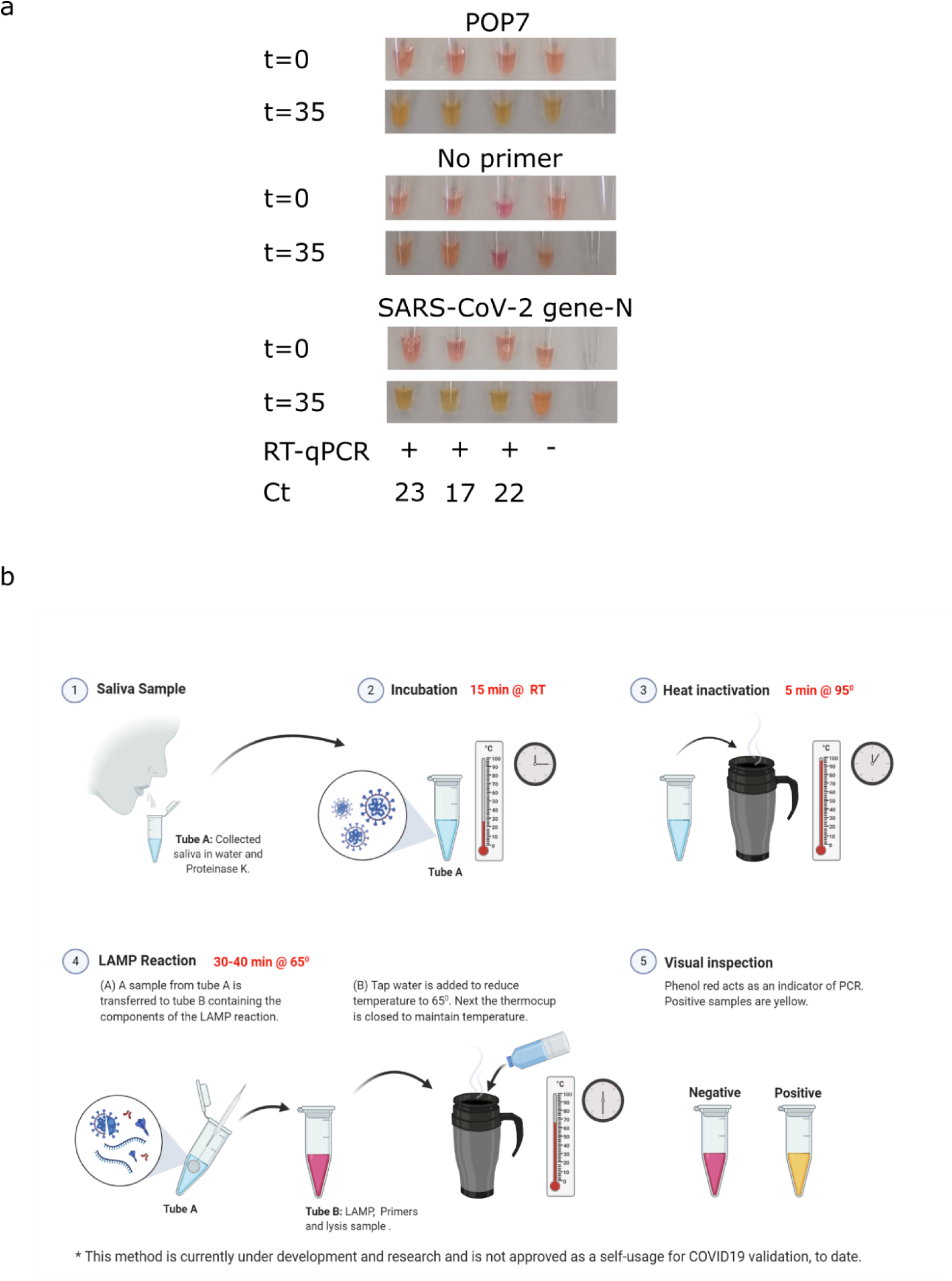
Applying the RT-LAMP protocol on saliva samples. **a**, RT-LAMP tests on saliva from 4 volunteers. Each tube represents one tested volunteer. Results of t=0 and t=35 are shown. Upper panel, RT-LAMP reaction using POP7 primers as a positive control. Middle panel, RT-LAMP reaction with no primers control. Lower panel, RT-LAMP reaction using SARS-CoV-2 gene N primers. The same samples were analyzed by the conventional hospital RT-qPCR protocol. The RT-qPCR results and Ct values are shown under the relevant samples. **b**, Graphical illustration of the potential of RT-LAMP protocol to perform self-saliva testing.

Due to the isothermal property of the reaction and the fact that the test is detectable in saliva samples, the reaction can potentially be performed in any constant heat source. Self-sampling can be done in a thermal-cup as shown in (Fig. 3b).

## Discussion

RT-LAMP is a rapid and simple method to detect purified RNA of SARS-CoV-2 ^[4]^. In this study, we were interested to test its potential as a direct detection method for viral presence in suspected COVID-19 patients. To do so, we adjusted and validated the RT-LAMP protocol for direct detection from clinical diagnostic patient swab samples. Our experiments were performed side-by-side to the standard RT-qPCR method at the hospital. We compared RT-qPCR Ct values to the RT-LAMP results of more than 180 different patients. Upon calibration, this direct RT-LAMP method successfully detected patients with medium to high viral loads, while yielding very few false-positives.

We demonstrate the optimal protocol for immediate off-the-shelf use of RT-LAMP on COVID-19 patients in the current pandemic. This protocol does not include RNA purification step. Besides a constant heat source, (e.g. a thermal mug), no sophisticated equipment is required. This protocol takes about an hour from sampling to detection, with very few reagents, and can potentially be performed by non-professionals or self-performed. These features allow its implementation around the globe, including in rural areas.

The next development steps required before clinical application are further adjustment of the protocol on saliva samples, tested on a large cohort, and compared to the standard method. Upon such further development, such a SARS-CoV-2 detection method can be applied as a surveillance tool for sampling larger populations of the community. Its simplicity, availability of products and low cost, makes it easy to continuously monitor suspected subjects. With further development, this method can be applied to medical clinics, points of entry, nursing homes, workplaces, etc. Importantly, this method can be easily adjusted to other emerging pathogens as well.

## Methods

### Samples collection

Swabs from both throat and nose were previously collected to one tube by healthcare providers and sent to the Virology laboratory at the Rambam Health Care Campus, Haifa, Israel. The swabs were stored in 1-2 ml of Universal Transport Medium (UTM). Saliva samples were self-collected directly into sterile cups, and kept at 4 °C until tested.

### Quantitative reverse transcription PCR (RT-qPCR)

Viral RNA was extracted by either of three automated nucleic acid extraction systems: (1) easyMAG ® / EMAG (Biomeriuex); (2) magLEAD® 5bL (Precision System Science); (3) MagEx (STARlet) using the following protocols: (1) 2 ml lysis buffer, 0.5 ml sample and 50 µl elution buffer; (2) 270 µl lysis buffer, 130 µl sample and 50 µl elution buffer; (3) 300 µl lysis buffer, 400 µl sample and 50 µl elution buffer, respectively. Following viral RNA extraction, RT-qPCR was performed using either of two commercial kits: (1) Allplex™ 2019-nCoV (Seegene); (2) Real-Time Fluorescent RT-PCR Kit for Detecting SARS-2019-nCoV (BGI), according to manufacturer’s instructions. Additional RT-qPCR reaction mix was created manually, using custom made primers (Primers table) as follows: The probe IC was synthesized with a 5’ FAM/CY5 Fluorophore moiety and 3’ ZEN/IBFQ quencher. Each of the manual reactions was assembled in a total volume of 25 µl. 12.5µl 2X Ag-Path One-step mix (Ambion), 1 µl primers, 1µl of Reverse Transcriptase enzyme, 5 µl of the sample’s RNA extracted previously and H_2_O to a final volume of 25 µl. All RT-qPCR reactions were executed in either of two systems: (1) Quantstudio (Thermo Fisher Scientific Inc.) or (2) CFX96 RealTime PCR machines (Bio-Rad) under the following conditions: 30 minutes at 50 °C, 10 minutes at 95 °C and 45 cycles of two-step incubations: (1) 15 seconds at 95 °C; (2) 30 seconds at 55 °C. Fluorescence was measured during step 2 of each cycle.

### Colorimetric RT-LAMP Reaction

5µl of UTM samples were diluted in 40 µl of DNase RNase free water (Biological Industries, 01-869-1B) and 2 µl of Proteinase K (1.22 mg/ml final concentration) (Seegene, 744300.4.UC384). Samples were incubated at room temperature for 15 minutes. Inactivation of proteinase K was obtained by incubating the reaction tubes at 95 °C for 5 minutes. Next, colorimetric RT-LAMP reaction was performed in a total volume of 20 µl per reaction using 10 µl WarmStart® Colorimetric LAMP 2X Master Mix (New England BioLabs Inc., M1800), 2 µl primers mix (see Primers Table), 1 µl Guanidine hydrochloride (Sigma, G4505), to a final concentration of 40 mM, and 7 µl of the inactivated sample. The reaction was then incubated for 30-40 minutes at 65 °C. The proteinase K inactivation step and the reaction step were performed in a closed lid heating-block or, for a proof of concept, in a thermos-cup, using warm water. In the case of a thermos-cup, temperature was monitored by a simple thermometer. After 20 minutes of incubation, samples were monitored for color change every 5 minutes, by the naked eye, until reaching 40 min incubation. Samples were considered negative if the original pink color of the phenol red was maintained and positive if the phenol red color turned orange-yellow.

(for the calibration process, reactions were performed with or without proteinase K and guanidine hydrochloride).

**Primers table:**

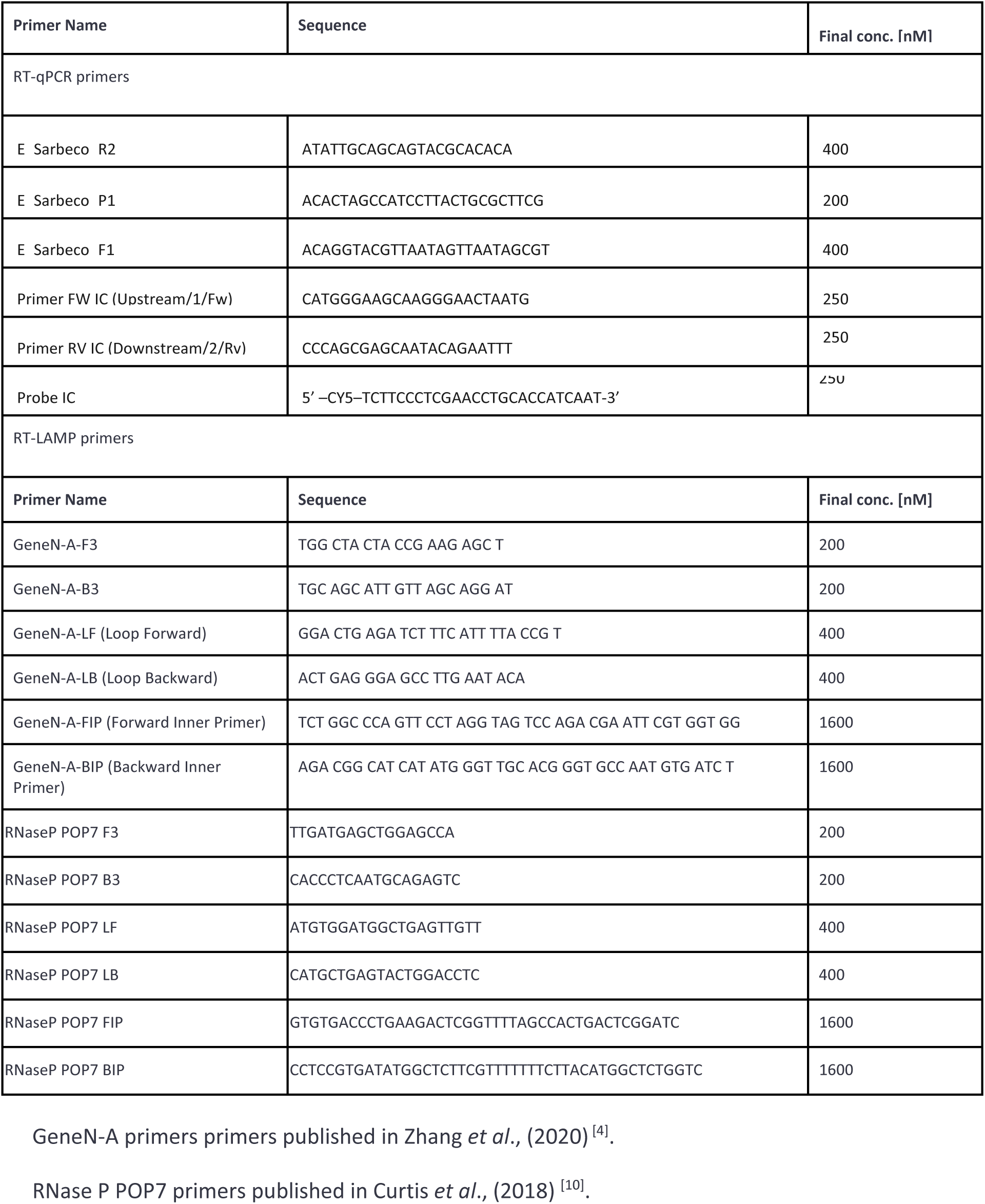

### Statistical analysis

TPR (true positive rate), TNR (true negative rate), FPR (false positive rate), FNR (false negative rate) were calculated according to the following equations: TPR= TP/(TP+FN). TNR=TN/(FP+TN). FNR=FN/(TP+FN). FPR=FP/(FP+TN). TP: total number of true positives. TN: total number of true negatives. TN: total number of true negatives. FN: total number of false negatives.

### Ethical approval

This study was granted exemption from IRB approval of the Rambam Health Care Campus for use of deidentified COVID-19 tests performed for the purpose of the standard testing, and for 4 volunteers.

## Data Availability

All data is available.

## Acknowledgements

We thank the Geva-Zatorsky lab for fruitful discussions and contributions, and the virology team at RAMBAM hospital for hosting us. We would also like to thank Dr. Rich Roberts and Dr. Nathan Tanner for their valuable support, as well as Prof. Yehuda Chowers, Dr. Ronit Almog, Dr. Yuval Geffen, Dr. Dani Zvi-Bar and Prof. Oded Lewinson for their help.

This work was supported by the Technion Integrated Cancer Center, the Technion - Israel Institute of Technology, “Keren Hanasi”, Alon Fellowship for Outstanding Young Researchers, Horev Fellow (Taub Foundation). HH is supported by the Leonard and Diane Sherman Interdisciplinary Graduate School Fellowship. NGZ is an Azrieli Global Scholar at the Canadian Institute for advanced research (CIFAR).

## Conflict of interest

The authors declare no conflict of interest

## Notes

### Competing Interest Statement

The authors have declared no competing interest.

